# Study design and rationale of Boxed-Breathing-Heart: a translational ex vivo study evaluating virus-mediated gene delivery for gene therapy in normothermic machine-perfused human hearts

**DOI:** 10.64898/2026.07.13.26357866

**Authors:** Isabel Branzei, Ali Amr, Kleopatra Rapti, Laura Schraft, Dominik Lindenhofer, Albrecht Leo, Gabriele Romano, Farbod Sedaghat-Hamedani, Christoph Reich, Jan Koelemen, Jan Haas, Ana Muñoz-Verdú, Jan Beckendorf, Philipp Schlegel, Willem H. te Gussinklo, Anna Meyer, Rawa Arif, Matthias Karck, Norbert Frey, Lars M. Steinmetz, Dirk Grimm, Benjamin Meder

## Abstract

Research on targeted genetic therapies for myocardial diseases, such as cardiomyopathies, currently focuses on (r)AAVs as the delivery method. Despite substantial efforts and advances in animal trials, predicting biodistribution and transduction efficacy in human tissue remains challenging due to interspecies differences in tissue tropism and the difficulty of accurately assessing alternative delivery routes and vector differences. The application in humans has also proven challenging, in part due to severe adverse events associated with systemic administration of (r)AAVs. This necessitates implementing alternative trial designs and stringent evaluation methods that minimize harm or risk to patients. Applying a predesigned vector carrying a gene-editing tool to a normothermic machine-perfused living beating heart in an ex vivo setting could overcome conventional obstacles and limitations. This can serve as a basis for safe and effective gene-therapy testing and assist in evaluating effects at the molecular level.

Boxed-Breathing-Heart is a translational trial assessing the feasibility of ex vivo gene editing and gene translation in normothermic machine-perfused human hearts. Human hearts explanted from cardiomyopathy patients undergoing heart transplantation are donated for research and immediately placed in an Organ Care System, where they are surgically connected. The viability of the heart is maintained through normothermic perfusion of system solutions and donor blood. A predesigned AAV containing a CRISPR-Cas system is infused into the circulation and dispersed throughout the tissue via coronary perfusion. The changes at the cellular and molecular levels are assessed continuously via frequent sequential myocardial biopsies. Furthermore, after the pre-planned 72-hour perfusion, the heart is sectioned and analyzed using spatial and single-cell omics. The aim is to provide a proof-of-concept for genetic therapeutic options delivered to the human heart via AAV in an ex vivo perfusion setup.

In summary, Boxed-Breathing-Heart provides an ex vivo translational platform for evaluating targeted cardiac gene therapies, enabling molecular analysis directly in human hearts and accelerating clinical translation without posing risks to patients.

## Background and study rationale

Current cardiovascular research is shifting its focus towards more personalized therapies targeting the genetic cause of myocardial diseases (1). This trend can be observed across different fields but is especially relevant for monogenetic and hereditary disorders such as cardiomyopathies, since conventional therapies largely focus on symptom relief based on a “one-size fits all” principle (2). Gene therapy has gained significant attention over the past decade and represents the next logical step in the paradigm change towards personalized precision medicine (3).

Recombinant adeno-associated viruses (rAAV) are the primary delivery strategy for these emerging technologies and are used in contemporary trials due to their favorable safety profile and varying tissue tropisms (4, 5). To analyze tissue tropism, extensive testing is conducted in vitro and in vivo (6). Nonetheless, the clinical translation of these technologies is limited by serious setbacks, including adverse events and fatalities in early- and late-phase trials (7). These limitations emphasize the risks associated with direct systemic AAV delivery to humans, necessitating alternative testing strategies (8).

While experiments on human induced pluripotent stem cell-derived cardiomyocytes (hiPSC-CMs) can demonstrate molecular effects and cellular efficiency, they fail to recreate the multicellular, three-dimensional architecture of human tissue (9, 10). Tissue slices, as a new technology, capture this composition (11). Even though the three-dimensional architecture is maintained during the cutting process, the diffusion barrier provided by the tissue vessels is not preserved. Animal models, including large-animal trials in pigs, can reproduce these anatomical details but differ in their genetic backgrounds. Since AAVs exhibit varying interspecies tissue tropisms, these models are also limited in their ability to address human-like conditions (12, 13). As a result, this leaves a research gap in evaluating delivery routes and vector designs under clinically relevant human genetic conditions.

The project Boxed-Breathing-Heart addresses this translational aspect by providing a platform that enables patient-relevant assessment of gene therapies at an intact-organ level. The surgical connection of a diseased heart to a machine perfusion system facilitates the use of gene therapy vectors in normothermic, living human hearts ex vivo. This trial evaluates biodistribution, transduction efficiency, and molecular effects in a controlled environment, allowing comparisons of delivery strategies and vector variants. By bridging the gap between preclinical models and clinical application in patients, this platform accelerates the safe and effective translation of personalized cardiac gene therapies.

## Objectives

The primary objective of the study is to develop a methodology that facilitates conducting gene-therapy experiments while safeguarding the well-being of vulnerable patients. The project is based on the mechanical perfusion of explanted, diseased human hearts utilizing an Organ Care System (OCS). This promises to represent the most accurate approximation to in vivo research attainable.

Patients who are undergoing heart transplantation can donate their sick heart for research purposes after receiving a donor heart. This approach has previously not been viable for time-consuming experiments, such as gene therapy, due to time restrictions related to ischemia (14, 15). Moreover, the limited time frame (up to 4 hours) could only be achieved by inducing hypothermia, thereby minimizing cellular myocyte metabolism to an absolute minimum (16, 17). Both prerequisites render the conventional use of explanted diseased hearts for gene-editing research as non-viable.

Normothermic machine perfusion is an ideal system for donor organ management, prolonging the survival of the beating heart ex vivo, thereby providing researchers with sufficient time to perform time-consuming experiments on living hearts without endangering the patient (18). The device used at the University of Heidelberg for heart transplantation is the TransMedics Organ Care System. This project aims to establish OCS systems as a platform that facilitate research on beating, normothermic, diseased human hearts.

The objective of the study is to utilize this platform to establish the foundational framework for implementing gene therapy using gene transfer, base editing, and other CRISPR-Cas technologies in living hearts. At first, a pathogenic variant will be introduced into myocardial tissue via an AAV9 vector. Subsequent experiments will focus on the targeted correction of pre-identified, patient-specific variants through reversal using a predesigned mutation-specific CRISPR-Cas system.

## Study design

AAV9, a viral vector with high cardiac tropism, is administered to the machine-perfused sick human heart (19, 20). Upon systemic delivery into the OCS or direct injection into the coronary arteries, the virus disseminates throughout the closed circulation and ultimately reaches the cardiomyocytes (Fig. 1). Regarding the AAV genome, regulatory elements and the gene cassette can be designed to tailor the rAAV to its intended applications, which aims to deliver a therapeutic gene cassette into the host cell, enabling its retention and functional expression, typically via episomal persistence, to compensate for the underlying genetic defect (21-26).

**Figure 1:**
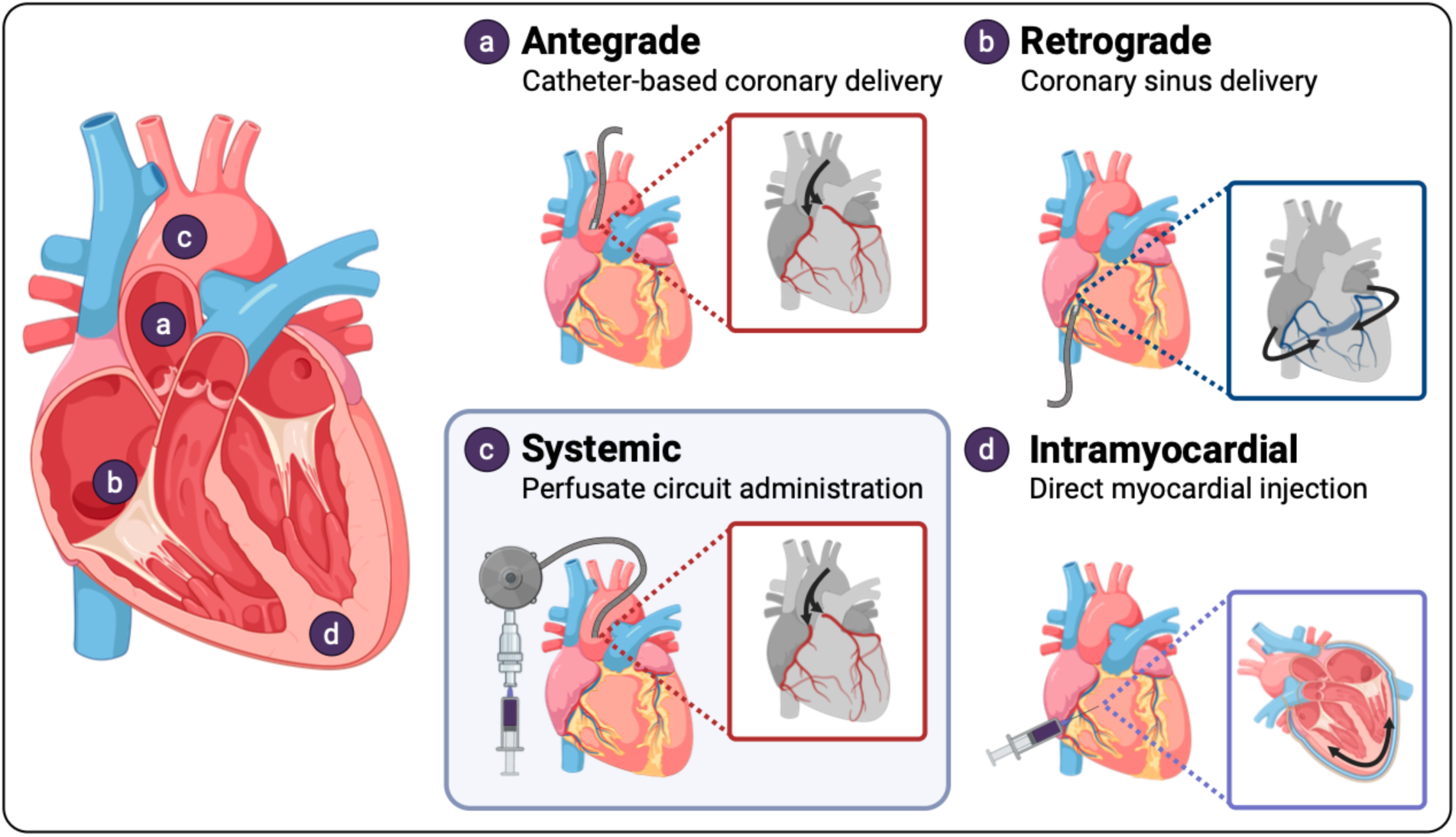
Contemporary AAV delivery routes. (a) Antegrade coronary delivery via catheterization of the aortic root. (b) Retrograde infusion via the coronary sinus. (c) Systemic administration to the perfusate via infusion into the reservoir or access via the system’s built-in ports, as used during the Boxed-Breathing-heart trial. (d) Direct intramyocardial delivery by injection into the ventricular myocardium.

Various factors must be evaluated regarding the virus itself, including the optimal application method to achieve favorable results (27-29). In the latest clinical studies, several application methods have been described, including antegrade, retrograde, systemic, and intramuscular deliveries (Fig. 1). In the proposed OCS platform, all the previously mentioned delivery methods can be compared, even within a single experiment, by tagging the delivered gene constructs for each application method. The distribution of the virus within myocardial tissue and its effects on the myocytes will also be comprehensively assessed and analyzed using sequential myocardial biopsies obtained via the surgically connected cannulas. The analyses will be performed using single-cell sequencing, which will provide insights into viral myocardial uptake, tissue distribution, and potential myocyte toxicity (30-32). Upon completion of the pre-planned 72-hour perfusion period, comprehensive histopathological evaluation will be performed after careful sectioning of the heart, followed by spatial and single-cell omics analyses.

## Organ source and selection

The Boxed-Breathing-Heart platform is an innovative concept that integrates human tissue in an ex vivo setting. Given the novelty of the project, an interdisciplinary team, including leading clinicians, molecular biologists, geneticists, heart surgeons, and bioinformaticians, conceptualized an ethical framework for the project. Special care was taken to ensure that the framework was in conformity with good clinical practice and in line with the Declaration of Helsinki. The well-being of all participating patients was consistently regarded as a primary concern. All patients will be required to provide written informed consent prior to being recruited. An application to the ethics committee at the University Hospital Heidelberg was submitted, and approval was granted on 11.04.2023 (Ethics approval number S-703/2022). Following the authorization, a logistical concept was established to ensure optimal coordination among the interdisciplinary team, prompt connection of the heart following explantation and the on-call availability of all contributing team members.

Patients aged ≥18 years listed for heart transplantation at the University Hospital Heidelberg will be invited to participate in the study. With their consent, the diseased heart that would otherwise be discarded can be utilized for the study. The individual anatomy will be assessed on a case-by-case basis using available patient scans. Since patients on the transplant high-urgency list undergo thorough analysis and multiple imaging sequences are available, no additional imaging will be conducted for this study. Patients will be deemed ineligible if their hearts cannot be connected to or perfused by the system, including but not limited to cases with extensive prior cardiac surgery or significant aortic regurgitation. No analysis of AAV-neutralizing antibodies will be conducted for the patient whose heart is donated, as the local tissue antibody levels are considered too insignificant to affect the trial (33, 34).

## Machine perfusion system

The OCS is a machine perfusion system designed to replicate the physiological conditions of the human body, thereby ensuring the adequate perfusion and oxygenation of the explanted heart. Blood and perfusion solutions are circulated through the system, propelled by a pulsatile pump. The gas exchanger continuously supplies fresh oxygen, while a warmer regulates the temperature to maintain a normothermic state reflective of human physiological conditions (35-38).

To facilitate oversight of this process, probes and vital parameters are monitored in real-time. A blood gas analyzer is used to assess blood gases, electrolytes, and lactate concentrations throughout the procedure. Electrolytes, particularly calcium and glucose, may require replenishment during the process and are monitored and adjusted accordingly. At the same time, the external access ports enable the introduction of catheters, probes, and additional monitoring instruments into the aorta. This capability enables tailored monitoring of the perfused heart and supports potential interventions. Such interventions may include the administration of fluids via infusion ports, while heart defibrillation using conventional methods is also feasible (38, 39).

In addition to the system solutions, blood is essential for perfusion. The optimal volume required ranges from 1200 to 1500 milliliters. Typically, this blood is sourced from the brain-dead patient whose heart is explanted for donation (38). In this project, the heart utilized has been explanted from a patient receiving a donor’s heart. Consequently, acquiring blood from the cardiac patient is not a viable option. Instead, the blood supply is sourced from blood donors taking part in this project, ensuring compatibility between the blood and the donated heart.

Recent studies have shown that the OCS heart system can successfully maintain the donor heart’s viability for several hours (36). Given that the OCS is not routinely used for extended periods, the perfusion protocol is altered for the project. A key aspect of the revised setup involves periodically removing metabolites that build up over time by continuously replacing a small portion of the perfusate (40). In contrast to contemporary studies conducted in the last five years on OCS machines, this project uses diseased hearts from patients diagnosed with terminal heart failure. Several aspects could play a significant role in determining the duration of viability, ranging from microcirculatory dysfunction to valvular impairment. The project also investigates the maximum duration for which a diseased heart can remain viable and survive under normothermic mechanical perfusion, thereby making this setup applicable to future research.

## Vector characteristics and administration during perfusion

The information encoded within the virus vector is subject to modification. A gene-editing tool based on a base editor, or the CRISPR-Cas system, can be integrated into the cassette (41). When the virus infiltrates the cell, the encoded information is simultaneously conveyed (42). During the first phase, it is imperative to ensure that the system is effective and efficient. This can be demonstrated through a straightforward alteration to the target cells, which minimizes the likelihood of complications. Consequently, during the initial application of this system, base editing will be performed to introduce a pathogenic RBM20 S637G variant (Fig. 2). Future steps will include the possibility of correcting a known disease-causing variant.

**Figure 2:**
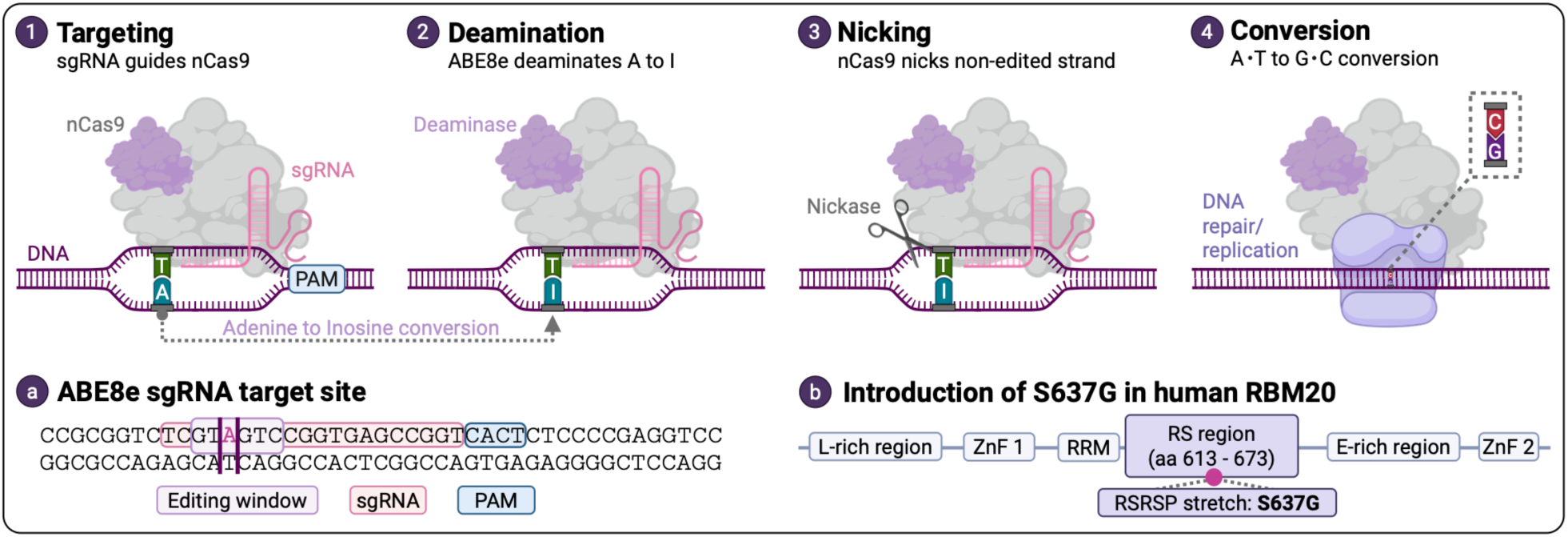
ABE8e-mediated introduction of the pathogenic RBM20 S637G variant. Panels 1-4 illustrate a schematic overview of adenine base editing. (1) Cas9 recognizes the protospacer adjacent motif (PAM) while the single-guide RNA (sgRNA) directs the complex to the complementary target sequence. (2) ABE8e deaminates adenine (A) to inosine (I) within the editing window. (3) nCas9 introduces a nick in the non-edited DNA strand, resulting in a higher likelihood of using the edited strand during repair and replication. (4) During subsequent DNA repair and replication, inosine is interpreted as guanine, leading to the replacement of A/T with G/C. (a) Sequence for the target site in RBM20 showing the sgRNA, PAM sequence, and editing window. The intended target site for the adenine deamination is highlighted (II). (b) Schematic domain organization of the human RBM20 protein structure. For the nuclear localization of RBM20, RRM and RS (especially RSRSP stretch) are contributing. Adapted from Lennermann et al. (2020) (47) Abbreviations: ABE (adenine base editor), L-rich region (leucine-rich region), ZnF (zinc finger domain), RRM (RNA recognition motif), E-rich region (glutamate-rich region).

Intracoronary administration via catheterization and direct systemic administration are both possible (Fig. 1). In the Boxed Breathing heart study, systemic delivery will be achieved through the system’s built-in application port, allowing the viral mixture to circulate continuously (38). This method simulates repetitive intracoronary administration due to the lack of accumulation in liver tissue. The predesigned vector circulates through the machine and repeatedly reaches the myocardial tissue via perfusion of the coronary arteries. It thereby reflects the current practice of intracoronary application for cardiac genetic therapy, similar to what’s used in modern human AAV studies (43-46).

## Tissue and sample collection

The clinical analysis of the tissue and its alterations during extended perfusion is thoroughly planned (Fig. 3). Major cardiac edema can be visually assessed by increased tissue volume and by measuring cardiac weight before and after perfusion (48-50). Contractility is likely to decrease during prolonged perfusion, necessitating adjustments to the flow parameters in the OCS system (51, 52). Arrhythmias may occur during perfusion, requiring pacing or defibrillation of the heart (38). In addition to these tissue-level changes, enzyme alterations can be analyzed to assess heart viability. Rising lactate and cardiac enzymes, such as NT-proBNP and troponin T, are expected and will be monitored during perfusion (53, 54). These parameters help determine changes in heart tissue metabolism during prolonged perfusion, indicate how long translational experiments can be performed with this system in the future, and allow conclusions about the feasibility of ex vivo research on machine-perfused hearts. A rise in these parameters will also trigger perfusate exchange to ensure optimal metabolic and nutrient supply.

**Figure 3:**
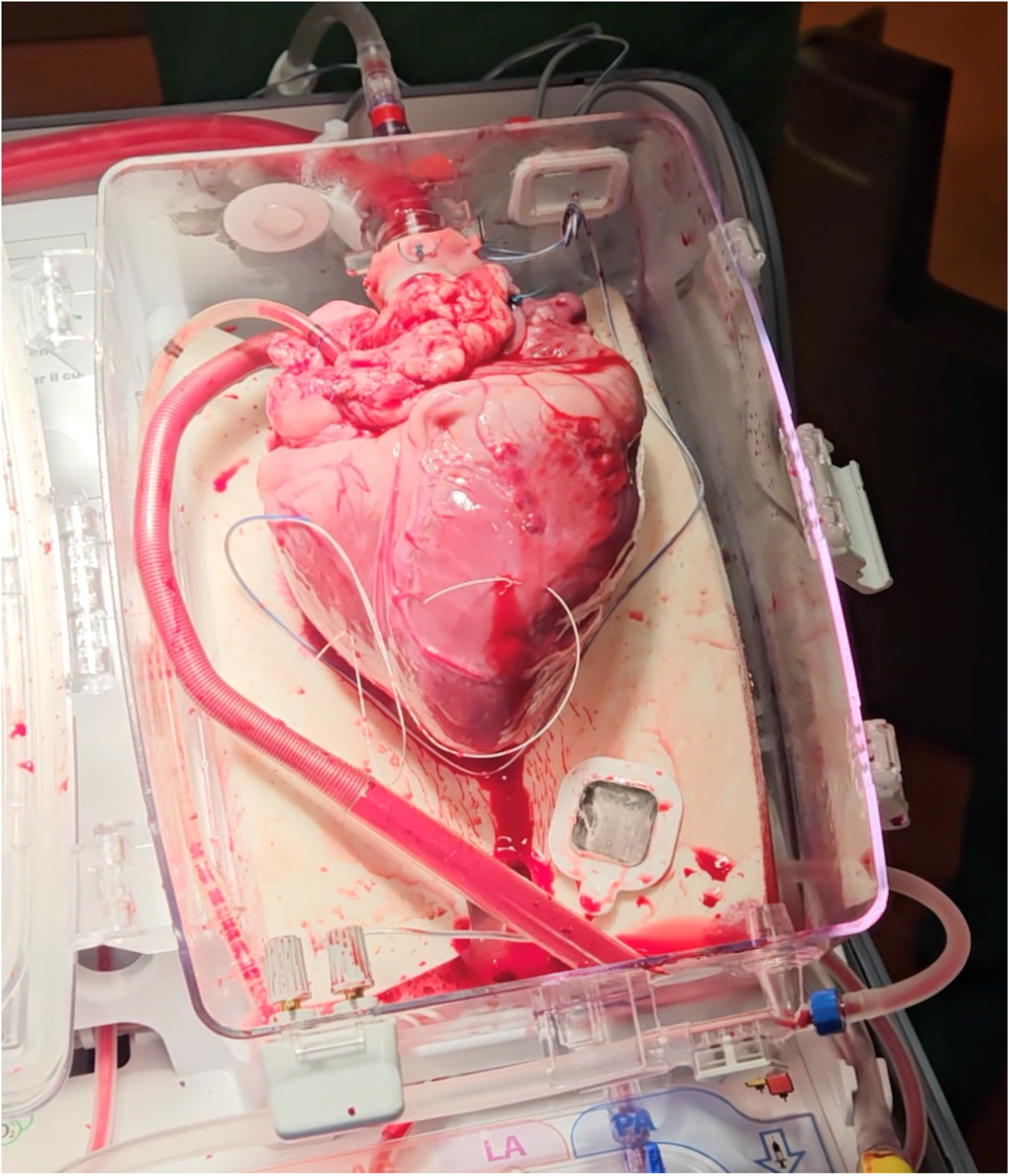
Heart situated in the heart chamber of the Organ Care System (OCS™ Heart, TransMedics Inc., Andover, MA, USA) during normothermic ex vivo perfusion.

Changes at the molecular level are monitored by obtaining myocardial biopsies at regular time intervals of every 6 hours. Tissue samples can be repeatedly obtained using a standard bioptome inserted through the system’s ports (Fig. 3). This setup, with sequential biopsies, is a first-of-its-kind approach for providing real-time insights into genetic alterations in human tissue based on single-cell sequencing. It allows observation of the timestamps at which integration occurs and when translation of the gene product can be expected. After perfusion for a pre-planned 72-hour period, the heart is sectioned for further analysis of vector distribution. By specifically labeling the slices, it is possible to assess the distribution of the virus and the genetic modifications it conveys to different anatomical localizations within the tissue. This method reveals the percentage of infiltrated myocardial cells and the biodistribution within the heart using spatial omics. Since direct application to humans makes numerous biopsies and heart-slicing after administration impossible, this setup offers great advantages over contemporary trials.

Arterial and venous ports allow administration of medication and continuous blood gas analysis. Direct access to the heart is feasible throughout perfusion, enabling interventions when required. Myocardial contractility can be assessed during perfusion by echocardiography.

## Outcome measures

The study is observational and descriptive and will evaluate edema, perfusion characteristics, and tissue changes to determine the suitability of prolonged normothermic machine perfusion for ex vivo research. At the molecular level, biopsies and tissue slices will undergo DNA, RNA, and protein extraction, followed by single-cell sequencing and spatial omics to assess gene product transcription and translation. This will allow for the analysis of biodistribution and can confirm that the setup provides a platform for gene therapy trials.

## Limitations

The study’s design limits the immune response. This is highly important and intentional to identify a method suitable for translational research without risking patient safety. However, when considering future applications in patients, it can only confirm the construct’s effectiveness, whereas the patients’ reaction to the viral vectors cannot be adequately evaluated (55). Circulating neutralizing antibodies also pose a significant challenge for virus-mediated gene transfer (56). Numerous studies and clinical trials measure pre-treatment antibody concentrations to exclude individuals with high levels (45, 57). During the trial, the blood used for perfusion will be pre-analyzed to determine whether these antibodies against the AAV9 vector are present (33). From these results, it is also possible to infer the role of antibodies in a controlled setting and draw conclusions for human application. Additionally, the liver is excluded from the trial. Typically, the liver significantly influences the distribution of the virus after systemic administration, as many viral vectors exhibit extensive liver tropism, thereby decreasing systemic concentrations (58, 59). This off-target effect and accumulation in liver tissue are not evaluated in this study.

## Translational outlook

The initial proof-of-concept phase, which incorporates implementing a pathogenic RBM20 variant using adenine base editing, focuses on laying the groundwork for gene editing in an ex vivo human heart (60). Going forward, we will explore the feasibility of reversing a disease-causing variant in cardiomyopathy patients undergoing heart transplantation. All cardiomyopathy patients on the high-urgency list for heart transplantation will undergo genetic screening. Patients who have been identified with an underlying monogenic cause for their cardiomyopathy will be recruited. Once a donor organ is allocated to the patient and the heart transplantation procedure is performed, we can obtain the diseased heart for the purpose of evaluating the potential for variant-specific gene therapy. A custom-designed, patient-specific CRISPR-Cas system aimed at reversing the disease-causing variant will be pre-developed and safely stored for every recruited patient. When a donor heart is assigned to one of these patients, the explanted diseased heart, previously genetically phenotyped, will be connected to the machine perfusion system at the time of transplantation. The viral vector carrying the specifically designed gene for this patient will be infused via the OCS System. Consequently, we will be able to offer insights into mitigating patient-specific genetic causes of cardiomyopathy. This proof-of-concept, addressing a patient-specific disease-causing variant, could serve as a foundation for future clinical trials. In addition, other vectors, tagged siRNAs and nanoparticles, can be tested as alternatives to virus-mediated gene transfer.

## Data Availability

All data produced in the present study are available upon reasonable request to the authors

## Notes

### Competing Interest Statement

The authors have declared no competing interest.

### Author Declarations

Ethics Committee of the Medical Faculty of Heidelberg University, Heidelberg, Germany, gave approval for this work on 11.04.2023 (Ethics approval number S-703/2022).

### Funding

- Professorship of Precision Digital Health, University Hospital Heidelberg
- Apple Inc.
- Daiichi Sankyo
- AstraZeneca
- Novo Nordisk
- Bristol Myers Squibb
- Klaus-Tschira Foundation (Heidelberg, Germany)
- Informatics for Life (Heidelberg, Germany)
- German Center for Cardiovascular Research (DZHK, Berlin, Germany)
- Leducq Foundation (CASTT)
- German Research Foundation (CRC1550 Molecular Circuits)

